# A longitudinal analysis of pneumococcal vaccine serotypes in pneumonia patients in Germany

**DOI:** 10.1101/2021.10.07.21264682

**Authors:** Christina Bahrs, Miriam Kesselmeier, Martin Kolditz, Santiago Ewig, Gernot Rohde, Grit Barten-Neiner, Jan Rupp, Martin Witzenrath, Tobias Welte, Mathias W. Pletz, for the CAPNETZ Study Group

## Abstract

Recently, a 15-valent (PCV15) and a 20-valent pneumococcal conjugate vaccine (PCV20) have been licensed by the US Food and Drug Administration and are under evaluation by the European Medicines Agency. PCV15 contains all serotypes of the 13-valent conjugate vaccine (PCV13) plus serotype 22F and 33F and PCV20 includes PCV13 serotypes plus serotypes 8, 10A, 11A, 12F, 15B, 22F, 33F. We investigated pneumococcal serotype distribution, secular trends and proportion of pneumonia caused by serotypes included in PCV13, PCV15, PCV20, and the 23-valent pneumococcal polysaccharide vaccine (PPV23) among adult patients with all-cause community-acquired pneumonia (CAP) between 2013 and 2019. We applied logistic mixed regression modelling to assess annual trends. Urine samples from adult patients with CAP treated in the community or hospital in Germany and included in the CAPNETZ study, a prospective multi-centre cohort study, were analysed by two serotype-specific multiplex urinary antigen detection assays (UAD1/UAD2) at Pfizer’s Vaccines Research and Development Laboratory. UAD1 detects serotypes in PCV13, UAD2 detects additional serotypes in PCV20 plus serotypes 2, 9N, 17F and 20. Out of 1,831 patients screened, urine samples with a valid UAD test result were available for 1,343 patients (73.3%). Among those patients, 829 patients (61.7%) were male, 792 patients (59.0%) were aged ≥60 years, 1038 patients (77.3%) had at least one comorbidity and 1,204 patients (89.7%) were treated in the hospital. The overall proportion of vaccine-type pneumonia among all-cause CAP for PCV13, PCV15, PCV20 and PPV23 was 7.7% (n=103), 9.1% (n=122), 12.3% (n=165) and 13.3% (n=178). Over the entire observation period, we did not observe evidence for significant annual trends in pneumococcal vaccine serotype coverage against pneumonia in adults (PCV13: OR 0.94, 95% CI 0.83-1.05; PCV15: OR 0.93, 95% CI 0.84-1.03; PCV20: OR 0.95, 95% CI 0.86-1.04; PPV23: OR 0.99, 95% CI 0.90-1.08). In conclusion, our data show i) no decline of PCV13 serotypes in all-cause CAP between 2013-2019 mainly due to a persistently high proportion of serotype 3 suggesting no meaningful effect of childhood PCV13 vaccination on PCV13 coverage in pneumonia in adults during this time period and ii) that the gap in the coverage between PCV20 and PPV23 was small and did not increase over the entire observation time.

---

Pneumococcal infections are globally the most frequent vaccine-preventable cause of death [1], and community-acquired pneumonia (CAP) caused by *Streptococcus pneumoniae* is the main burden of pneumococcal disease in the elderly [2]. Since respiratory and blood cultures remain often negative in hospitalized patients with pneumococcal CAP due to prior antibiotic treatment, most cases are detected by the pneumococcal urinary antigen test (PUAT, BinaxNOW® *S. pneumoniae*) [2, 3]. As the PUAT does not allow serotype discrimination, data on serotype distribution in adult non-bacteremic pneumococcal CAP patients are sparse [4]. Pneumococcal conjugate vaccines (PCVs), which were primarily developed for vaccination of infants under 2 years of age, have significantly decreased invasive pneumococcal diseases worldwide in all age groups by herd protection effects [5, 6]. However, serotype replacement, i.e. replacement of vaccine serotypes by non-vaccine serotypes, has decreased the serotype coverage of PCVs over time [6, 7]. For Germany, we have described earlier the distribution of vaccine serotypes covered by the first but no longer available 7-valent conjugate vaccine and by the 13-valent conjugate vaccine (PCV13) between 2002 and 2016 in adult patients with CAP enrolled into the prospective multicentre study CAPNETZ [8, 9]. PCV7 was replaced by either the 10-valent conjugate vaccine (PCV10) or mainly PCV13 in the German infant vaccination program in 2010. However, PCV10 held the smallest market share of only 8% of pneumococcal vaccines in Germany in 2018 (https://www.fortunebusinessinsights.com/industry-reports/germany-pneumococcal-vaccines-market-101808 – last accessed 19^th^ October 2021). In adults, the German Standing Committee on Immunization (STIKO) recommends the 23-valent pneumococcal polysaccharide vaccine (PPV23) for all adults of 60 years and above and for all patients with defined chronic comorbidities predisposing to pneumococcal disease regardless of age. Moreover, since 2016, sequential vaccination with PCV13 followed by PPV23 is recommended for German adults at high risk for pneumococcal disease including individuals with immunosuppression, chronic liver disease, chronic kidney disease and individuals with cerebrospinal fluid leaks or cochlear implants [10]. Recently, a 15-valent (PCV15) and a 20-valent conjugate vaccine (PCV20) have been licensed for the adult indication by the US Food & Drug Administration and are under evaluation by the European Medicines Agency [11, 12]. PCV15 contains all serotypes of PCV13 plus serotype 22F and 33F and PCV 20 includes PCV13 serotypes plus serotypes 8, 10A, 11A, 12F, 15B, 22F, 33F.

The aim of the present study was to evaluate serotype distribution, secular trends and proportion of pneumonia caused by serotypes included in PCV13, PCV15, PCV20, and the 23-valent pneumococcal polysaccharide vaccine (PPV23) among adult patients with all-cause CAP between 2013 and 2019. All patients enrolled in the CAPNETZ study in Germany between January 1, 2013 and December 31, 2019 with an available urine sample were included in the analysis. The CAPNETZ study (German Clinical Trials Register: DRKS00005274; approval number of leading Ethics Committee “Medical Faculty of Otto-von-Guericke-University Magdeburg” No. 104/01, see acknowledgment or www.capnetz.de for participating centres) is a prospective observational multi-centre cohort study of CAP-patients treated in the hospital or in the outpatients setting. CAPNETZ inclusion criteria were age ≥18 years, radiologically-confirmed pneumonia, and at least one of the following clinical findings: cough, purulent sputum, fever or focal chest sign on auscultation. Exclusion criteria were hospitalisation during 28 days preceding the study, immunosuppression and active tuberculosis [13]. All patients provided written informed consent prior enrolment to the study. Urine samples of enrolled patients were prospectively collected and immediately treated with 0.5M 1,4-Piperazinediethanesulfonic acid buffer (Boston BioProducts) to a final concentration of 25mM to stabilize respective polysaccharides. Two serotype-specific urine antigen detection (UAD) assays [14, 15] covering different serotypes on urine samples were performed and analysed at Pfizer’s Vaccines Research and Development Laboratory (Pearl River, NY, USA). The UAD assay is a limit assay that uses Luminex technology, with positivity cut-off limits (based on antigen concentrations read off a standard curve), established for each serotype using 400 control urine specimens collected from otherwise healthy adults without CAP. Using nonparametric tolerance intervals, the assay is set to achieve at least 97% specificity for each serotype. UAD1 covers PCV13 serotypes [14] and UAD2 covers 11 additional serotypes (the seven included in PCV20, i.e. ST8, ST10A, ST11A, ST12F, ST15B, ST22F, ST33F, and the four included in PPV23, i.e. ST2, ST9N, ST17F, ST20) [15]. UAD analyses were performed as described previously [14, 15]. Results were classified into “positive”, “indeterminate” (excluded from analysis) and “negative”. According to the recommendation of the German Standing Committee on Immunization (STIKO) for pneumococcal vaccination in adults, patients were classified as “at risk for pneumococcal disease” based on age ≥60 years or on the presence of at least one comorbidity regardless of age [10]. We quantified the distributions of pneumococcal vaccine serotypes of PCV13, PCV15, PCV20, and PPV23 as absolute and relative frequencies (relative to the number of patients with information on the respective serotype). Furthermore, we applied logistic mixed regression modelling to assess annual trends (dependent variable: each of PCV13, PCV15, PCV20, PPV23 and serotype 3; independent variable: year of CAP acquisition; random effect (intercept): study centre; reported results: odds ratio (OR) with 95% confidence interval (CI)).

Out of 1,831 patients screened, urine samples with a valid UAD test result were available for 1,343 patients (73.3%) who were enrolled by 26 CAPNETZ centres distributed widely over Germany. Among those patients, 829 patients (61.7%) were male, 792 patients (59.0%) were aged ≥60 years, 1,038 patients (77.3%) had at least one comorbidity, 1,204 patients (89.7%) were treated in the hospital. Among the 1,108 patients at risk for pneumococcal disease, only 179 patients (16.2%) reported any pneumococcal vaccination within the last five years. In the overall study population during the study period 2013 to 2019, 183 of 1343 (13.6%) patients had a positive UAD1/2 test result. The most common vaccine serotypes were serotype 3 (n = 49; 3.7% of all-cause CAP), followed by serotype 8 (n=21; 1.6% of all-cause CAP), serotype 22F (n=13; 1.0% of all-cause CAP) and serotype 11A (n=11; 0.8% of all-cause CAP). As shown in table 1, the overall proportion of vaccine-type pneumonia among all-cause pneumonia for PCV13, PCV15, PCV20 and PPV23 was 7.7% (n=103), 9.1% (n=122), 12.3% (n=165) and 13.3% (n=178), respectively. When regarding only pneumococcal pneumonia diagnosed by conventional diagnostics (PUAT or blood culture; n=74), PCV13, PCV15, PCV20 and PPV23 coverage was 37.8% (n=28), 44.6% (n=33), 64.9% (n=48), and 66.2% (n=49), respectively. Bacteraemic pneumococcal CAP was detected in 19 (2.1%) of the 889 patients of whom blood cultures were obtained. Among them, bacteraemic pneumococcal CAP was caused by serotype 8 in four patients (21.1%), serotype 4 and serotype 7F in two patients (10.5%), and serotype 3, serotype 12F, serotype 14, serotype 20 as well as serotype 33F in one patient (5.3%) each. The coverage of PCV13, PCV15, PCV20, PPV23 in patients with bacteraemic CAP was 31.6% (n=6), 36.8% (n=7), 63.2% (n=12), and 68.4% (n=13). Over the entire observation period, we did not observe evidence for significant annual trends in pneumococcal vaccine serotype coverage (serotype 3: OR 0.95, 95% CI 0.81-1.10; PCV13: OR 0.94, 95% CI 0.83-1.05; PCV15: OR 0.93, 95% CI 0.84-1.03, PCV20: OR 0.95, 95% CI 0.86-1.04; PPV23: OR 0.99, 95% CI 0.90-1.08). Table 1 provides the serotype proportions of all-cause CAP for three time periods (2013-2014, 2015-2017 and 2018-2019) and the serotype proportion stratified by the above mentioned two STIKO classifications for patients “at risk” for pneumococcal disease (age ≥60 years or patients 18-59 years with ≥1 comorbidity). Serotype 3 was the most prevalent serotype in both patient subgroups, while the second most prevalent serotype was serotype 8 in patients 18-59 years with at-risk condition and serotype 11A in patients ≥60 years.

**Table 1.**
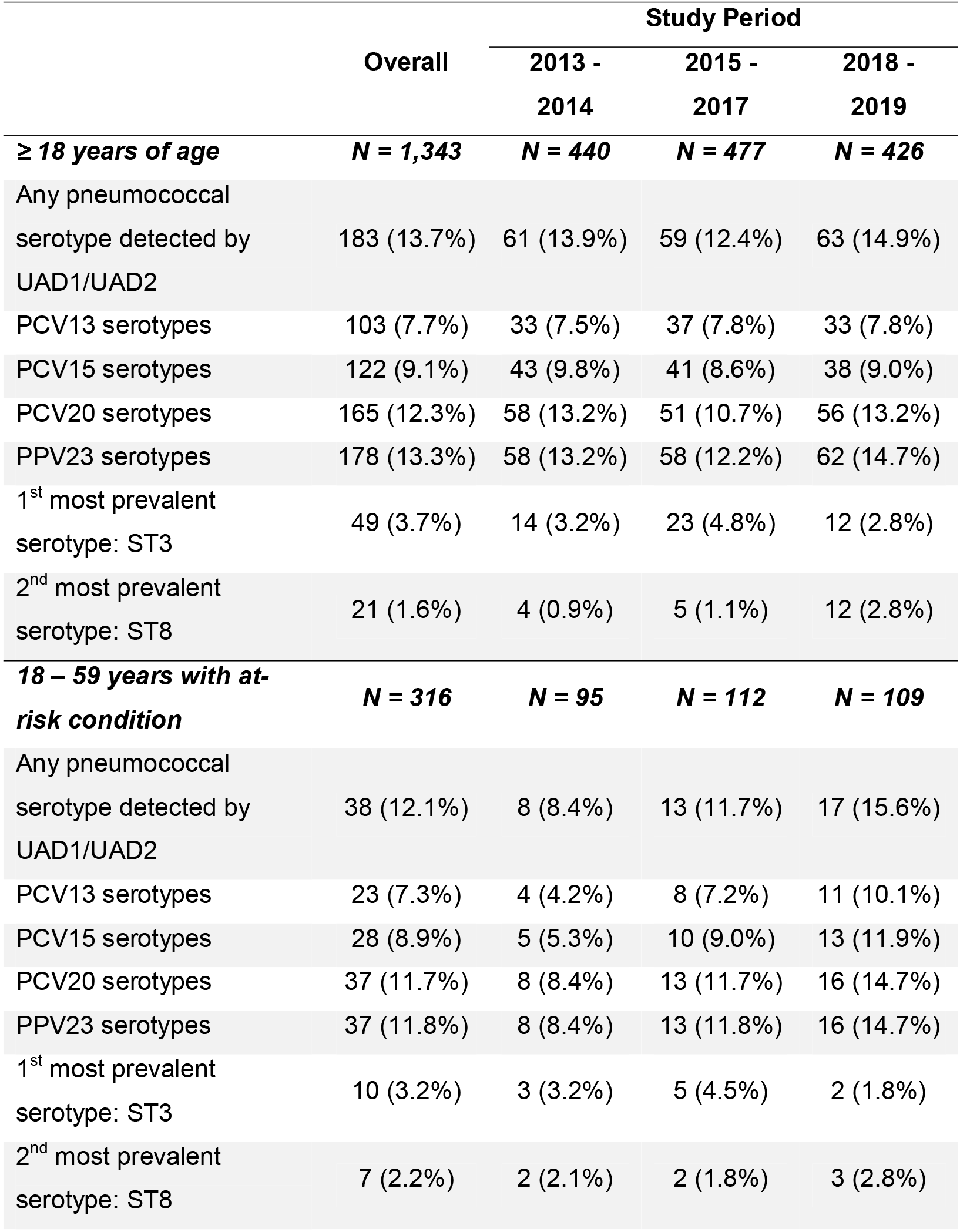
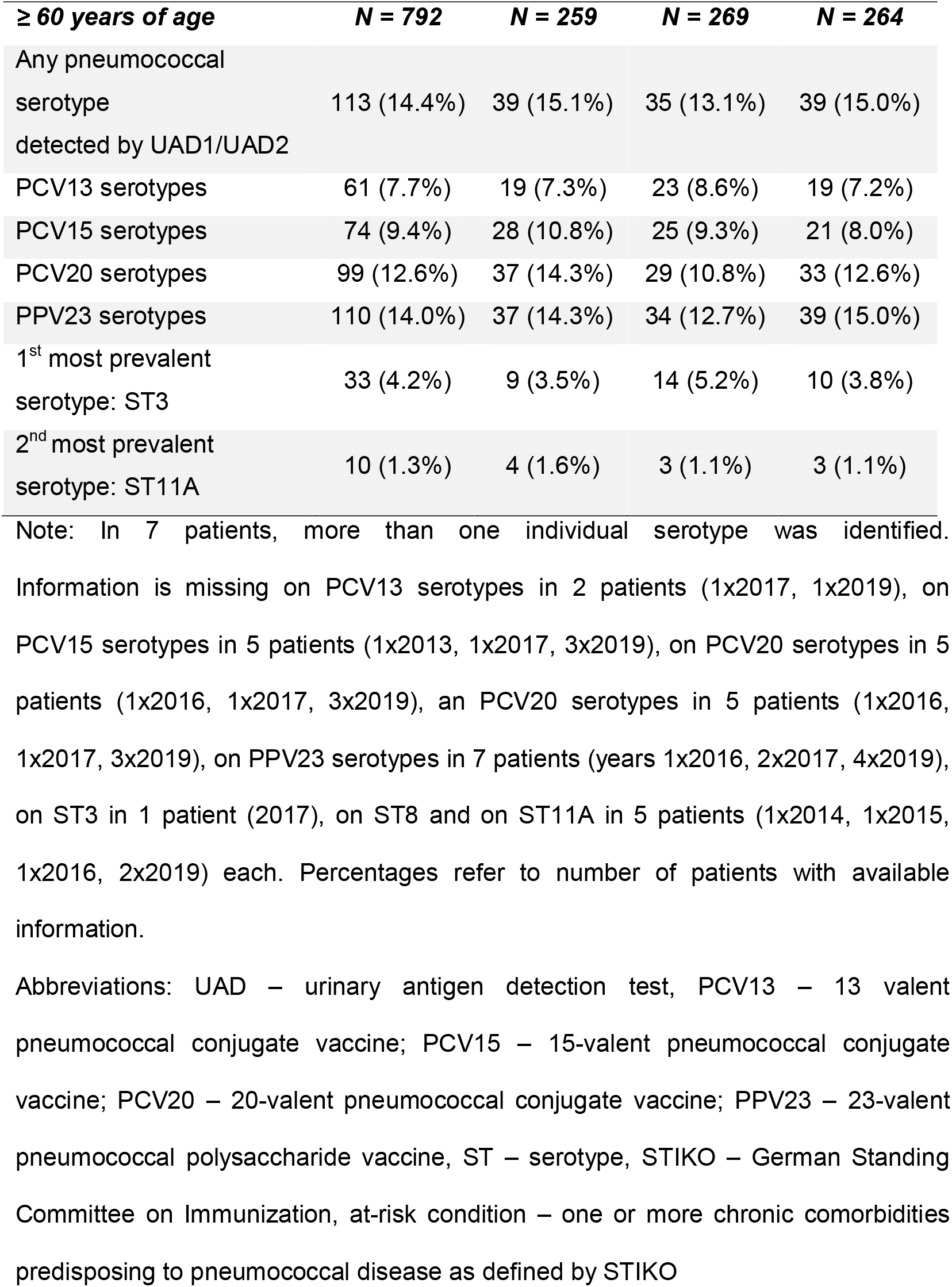
Distribution of pneumococcal serotypes aggregated by pneumococcal vaccine formulation in patients with radiologically-confirmed community-acquired pneumonia by UAD1/UAD2 by study period and in patient subgroups with STIKO recommendation for pneumococcal vaccination (individuals ≥60 years and individuals 18-59 years with at risk condition, i.e. ≥1 comorbidity)

In conclusion, PCV20 had a substantially higher coverage of all-cause CAP in adults compared to PCV13 (11.7% versus 7.3%) for age group 18-59 years with ≥1 comorbidity and 12.6% versus 7.7% for age group ≥60 years. Our data show i) no evidence of a decline of PCV13 serotypes in all-cause CAP between 2013-2019 mainly due to a persistently high proportion of serotype 3 suggesting no meaningful effect of childhood PCV13 vaccination on PCV13 coverage in pneumonia in adults during this time period and ii) that the gap in the coverage between PCV20 and PPV23 was small and did not increase over the entire observation time. The presented data may be of use for modelling impact of pneumococcal vaccines and may contribute to informed decision making of vaccination committees.

## Data Availability

All data produced in the present study are available upon reasonable request to the authors

## Funding

CAPNETZ was founded by a German Federal Ministry of Education and Research grant (01KI07145) 2001-2011. Since 2013 CAPNETZ was supported by the German Center for Lung Research (DZL): 2013-2015 Funding code 82DZL00204 and 2016-2020 Funding code 82DZL002A4. CB and MWP are partly supported by a grant of the Federal Ministry of Education and Research KliFo 2.0 (grant number 01KI1501). MW is supported by grants from the German Research Foundation, SFB-TR84 C6 and C9, SFB 1449 B2, by the German Ministry of Education and Research (BMBF) in the framework of the CAPSyS (01ZX1304B), CAPSyS-COVID (01ZX1604B), SYMPATH (01ZX1906A), PROVID (01KI20160A) P4C (16GW0141), MAPVAP (16GW0247), NUM-NAPKON (01KX2021), and by the Berlin Institute of Health (CM-COVID). This study was supported by an unrestricted grant from Pfizer.

## Author’s contribution and guarantor statement

All authors have made substantial contribution to the study design, data collection, analysis or interpretation, drafting the article and revising it critically for important intellectual content. All authors approved the final version to be submitted. MK, SE, GR, GB-N, JR, MW, TW, MWP designed the study, CB, MKe and MWP drafted the article, CB and MKe performed the statistical analysis. CB, MK, MKe, SE, GR, GB-N, JR, MW, TW, MWP contributed to the critical revisions, and final approval of the article.

## Potential conflict of interests

CB is a member of the scientific advisory board of GSK and reports personal fees from Pfizer for lectures and has received supports for attending meetings and travel, all outside the submitted work. MK reports personal fees from Berlin Chemie, Boehringer, Astrazeneca, Biotest, Novartis, GSK, Gilead, Pfizer and MSD and a research grant from Pfizer outside the submitted work. SE is member of the scientific advisory board of Pfizer. GR reports personal fees from Astrazeneca, Berlin Chemie, BMS, Pfizer, Boehringer Ingelheim, Solvay, Insmed, GSK, Essex Pharma, MSD, Grifols, Chiesi, Vertex, Roche, Takeda and Novartis for lectures including service on speakers’ bureaus outside the submitted work and/or consultancy during advisory board meetings and personal fees from GSK for travel accommodations/meeting expenses, outside the submitted work. As part of her activity as a member of the executive bodies, GB-N reports economic connections to the following diagnostic and pharmaceutical companies: ThermoFisher Scientific / BRAHMS, Alere Technologies GmbH, Merck Sharp & Dohme Corp., Pfizer Pharma GmbH, R-Biopharm AG and Helmut Hund GmbH. MW received personal fees from Astrazeneca, Bayer Health Care, Berlin Chemie, Biotest, Chiesi, Novartis, Teva, and research funding from Actelion, Bayer Health Care, Biotest, Boehringer Ingelheim, all unrelated to the current work. TW is the head of clinical studies and a member of the speakers’ bureau for Pfizer, Bio Merieux and Roche Diagnostics and is a consultant to Pfizer and MSD. MWP is a consultant to and a member of the speakers’ bureau for Bayer, MSD, Pfizer, meduptodate, Thermofisher and Novartis and has received research grants from Pfizer. JR and MKe have declared no conflict of interest.

## Acknowledgements

CAPNETZ is a multidisciplinary approach to better understand and treat patients with community-acquired pneumonia. Members of the CAPNETZ study group are: W. Knüppel (Department of Internal Medicine, Bad Arolsen Hospital); D. Stolz (Department of Pneumology, University Hospital Basel, Switzerland); W. Bauer (Central Emergency Admission / Medical Admission Ward, Charité-Universitätsmedizin Berlin); N. Suttorp, A. Mikolajewska, M. Witzenrath, (Medical Department, Division of Infectiology and Pneumonology Charité-Universitätsmedizin Berlin); S. Gläser, D. Thiemig (Department of Internal Medicine – Pneumology and Infectiology, Vivantes Hospital Neukölln, Berlin-Neukölln); C. Boesecke (Medical Clinic and Policlinic I – General Internal Medicine, University Hospital Bonn); M. Prediger (III. Medical Clinic, Carl-Thiem Hospital Cottbus); B. Schaaf, J. Kremling, D. Nickoleit-Bitzenberger (Pneumology, Infectiology and Internal Intensive Care Medicine, Medical Clinic Nord, Dortmund); M. Kolditz, B. Schulte-Hubbert, S. Langner (Medical Clinic I Department of Pneumology, University Hospital Dresden); C. Stephan (Medical Clinic II / Infectiology, University Hospital Frankfurt); G. Rohde, C. Bellinghausen (Medical Clinic I – Pneumology/Allergology, University Hospital Frankfurt); M. Panning (Institute of Virology, University Hospital Freiburg); C. Neurohr (Department of Pneumology and Respiratory Medicine, Clinic Schillerhöhe, Gerlingen); T. Welte, I. Pink (Department of Pneumology, Hannover Medical School, Hanover); G. Barten-Neiner, W. Kröner, M. Nawrocki, J. Naim, (CAPNETZ Office, Hannover); T. Illig, N. Klopp (Hannover Unified Biobank, Hannover Medical School); M. Pletz, B. Schleenvoigt, C. Bahrs (Institute of Infectious Diseases and Infection Control (IIMK), University Hospital Jena); C. Kroegel, A. Moeser (Clinic for Internal Medicine I, Department of Cardiology, Angiology, Pneumology, Internistic Intensive Medicine, University Hospital Jena); D. Drömann, P. Parschke, K. Franzen (Medical Clinic III, Pneumology/Infectiology, University Medical Center Schleswig-Holstein, Lübeck); J. Rupp, N. Käding (Department of Infectiology and Microbiology, University Hospital Schleswig-Holstein, Lübeck); G. Wesseling, K. Walraven, D. Braeken (Department of Respiratory Medicine, Maastricht University Medical Center, The Netherlands); C. Spinner (Department of Internal Medicine II, University Hospital rechts der Isar, Technical University of Munich, School of Medicine); A. Zaruchas (Medical Clinic-Pneumology, Brüderkrankenhaus St. Josef, Paderborn); M. Falcone, G. Tiseo (Department of Clinical and Experimental Medicine, Universita die Pisa, Italy); D. Heigener, I. Hering (Department of Pneumology, Agaplesion Diakonieklinikum Rotenburg); W. Albrich, F. Waldeck, F. Rassouli, S. Baldesberger (Department of Infectiology and Hospital Hygiene, Kantonsspital St. Gallen, Switzerland); S. Stenger (Institute for Medical Microbiology and Hygiene, University Hospital Ulm); M. Wallner (2mt Software, Ulm); H. Burgmann, L. Traby, L. Schubert (Department of Internal Medicine I, Division of Infectious Diseases and Tropical Medicine, Medical University of Vienna); and all study nurses.

## Abbreviation list

CAP: community-acquired pneumonia
CI: confidence interval
OR: odds ratio
PCV7: 7-valent conjugate vaccine
PCV10: 10-valent conjugate vaccine
PCV13: 13-valent conjugate vaccine
PCV15: 15-valent conjugate vaccine
PCV20: 20-valent conjugate vaccine
PPV23: 23-valent pneumococcal polysaccharide vaccine
STIKO: German Standing Committee on Immunization
UAD: serotype-specific multiplex urinary antigen detection assay

## Notes

### Competing Interest Statement

CB is a member of the scientific advisory board of GSK and reports personal fees from Pfizer for lectures and travel, all outside the submitted work. MK reports personal fees from Berlin Chemie, Boehringer, Astrazeneca, Biotest, Novartis, GSK, Gilead, Pfizer and MSD and a research grant from Pfizer outside the submitted work. SE is a member of the scientific advisory board of Pfizer. GR reports personal fees from Astrazeneca, Berlin Chemie, BMS, Pfizer, Boehringer Ingelheim, Solvay, Insmed, GSK, Essex Pharma, MSD, Grifols, Chiesi, Vertex, Roche, Takeda and Novartis for lectures including service on speakers' bureau outside the submitted work and/or consultancy during advisory board meedings and personal fees from GSK for travel accommodations/meeting expenses, outside the submitted work. As part of her activity as a member of the executive bodies, GB-N reports economic connections to the following diagnostic and pharmaceutical companies: ThermoFisher Scientific / BRAHMS, Alere Technologies GmbH, Merck Sharp and Dohme Corp., Pfizer Pharma GmbH, R-Biopharm AG and Helmut Hund GmbH. MW received personal fees from Astrazeneca, Bayer Health Care, Berlin Chemie, Biotest, Chiesi, Novartis, Teva, and research funding from Actelion, Bayer Health Care, Biotest, Boehringer Ingelheim, all unrelated to the current work. TW is the head of clinical studies and a member of the speakers' bureau for Pfizer, Bio Merieux and Roche Diagnostics and is a consultant to Pfizer and MSD. MWP is a consultant to and a member of the speakers' bureau for Bayer, MSD, Pfizer, meduptodate, Thermofisher and Novartis and has received research grants from Pfizer. JR and MKe have declared no conflict of interest.

### Author Declarations

The Ethics Committee Medical Faculty of Otto-von-Guericke-University Magdeburg gave ethical approval for this work

### Summary of Updates

The introduction and results were extended and the conclusion was adapted.

